# Outcome reporting from protocols of clinical trials of Coronavirus Disease 2019 (COVID-19): a review

**DOI:** 10.1101/2020.03.04.20031401

**Authors:** Ruijin Qiu, Xuxu Wei, Mengzhu Zhao, Changming Zhong, Chen Zhao, Jiayuan Hu, Min Li, Ya Huang, Songjie Han, Tianmai He, Jing Chen, Hongcai Shang

## Abstract

**Objectives:** To examine heterogeneity of outcomes in protocols of clinical trials of Coronavirus Disease 2019 (COVID-19) and to identify outcomes for prioritization in developing a core outcome set (COS) in this field.

**Design:** This study is a review.

**Data sources:** Databases of ICMJE-accepted clinical trial registry platform were searched on February 14, 2020.

**Eligibility Criteria:** Randomized controlled trials (RCTs) and non-RCTs of COVID-19 were considered. Conditions of patients include common type, severe type or critical type. Interventions include traditional Chinese medicine (TCM) and Western medicine. We excluded trials that for discharged patients, psychological intervention and complications of COVID-19.

**Data extraction and synthesis:** The general information and outcomes, outcome measurement instruments and measurement times were extracted. The results were analysed by descriptive analysis.

**Results:** 19 registry platforms were searched. A total of 97 protocols were included from 160 protocols. For protocols of TCM clinical trials, 76 outcomes from 16 outcome domains were reported, and almost half (34/76, 44.74%) of outcomes were reported only once; the most frequently reported outcome was time of SARS-CoV-2 RNA turns to negative. 27 (27/76, 35.53%) outcomes were provided one or more outcome measurement instruments. 10 outcomes were provided one or more measurement time frame. For protocols of western medicine clinical trials, 126 outcomes from 17 outcome domains were reported; almost half (62/126, 49.21%) of outcomes were reported only once; the most frequently reported outcome was proportion of patients with negative SARS-CoV-2. 27 outcomes were provided one or more outcome measurement instruments. 40 (40/126, 31.75%) outcomes were provided one or more measurement time frame.

**Conclusion:** Outcome reporting in protocols of clinical trials of COVID-19 is inconsistent. Thus, developing a core outcome set is necessary.

**Strengths and limitations of this study:**

1. This review is the first to describe variation in outcomes, outcome measurement instruments and outcome measurement time reporting in clinical trials for Coronavirus Disease 2019 (COVID-19).
2. All the database of ICMJE-accepted clinical trial registry platform were searched, and randomized controlled trials and observational studies were considered.
4. The aim of this review was to provide a list of outcomes for clinical trials of COVID-19, both interventions of Traditional Chinese Medicine and western medicine were considered.
5. When the searching was conducted, no clinical trials were registered by countries out of China, so all of included protocols were from China.

## INTRODUCTION

Since the severe acute respiratory syndrome coronavirus 2 (SARS-CoV-2) infection occurred in Wuhan, Hubei Province from December 2019, the disease, which was named as Coronavirus Disease 2019 (COVID-19) by World Health Organization (WHO) on February 12, 2020. According to the website of National Health Commission of the People’s Republic of China (NHC-PRC), 77,658 confirmed cases have been reported from all areas of China until 0 o’clock, February 25, 2020. 27,323 cured patients discharged, 2,663 patients died [1]. On the website of WHO showed that 2559 confirmed cases have been reported in 33 countries out of China, 34 patients died at 10AM CET, February 25 2020 [2].

However, there is still no specific medicine for COVID-19 now. In China, the government encourages traditional Chinese medicine (herbal medicine, moxibustion, Baduanjin, etc.) to take an important role in clinical practice. The NHC-PRC and National Administration of Traditional Chinese Medicine (NATCM) have released the Version 6.0 of Diagnosis and Treatment Guideline for COVID-19 (informal version) on February 18, 2020 [3]. The guideline recommended general therapy methods, such as oxygen support, or trying to use alpha-interferon, Lopinavir/Ritonavir, Ribavirin, Chloroquine phosphate, Abidol, etc. For severe and critical type of disease, high flow nasal catheter oxygen therapy, invasive or non-invasive mechanical ventilation, extracorporeal membrane oxygenation (ECMO), plasma of survivors, glucocorticoid, plasma exchange or according to the patients’ situation. The guideline also recommended TCM therapy methods, including herbal medicine formulas and proprietary Chinese medicine according to TCM syndromes, which are analyzed by clinical symptoms and signs through four methods of diagnosis: inspection, auscultation and olfaction, interrogation, and palpation.

At the same time, an increasing number of clinical trials are conducting. After searching some protocols of clinical trials from Chinese Clinical Trial Registry (ChiTCR) and ClinicalTrials.gov, we found that different researchers chose different outcomes. It is very important for clinical trials to provide evidence in treating COVID-19. However, the heterogeneity of outcomes make it impossible to conduct meta-analysis in the future, which may reduce the value of clinical trials and improve waste.

We are going to develop a core outcome set (COS) for clinical trials of COVID-19. The study have been registered in Core Outcome Measures in Effectiveness Trials (COMET) database [4]. The first clinical trial of COVID-19 was registered on January 23, 2020 [5]. When we registered the COS study, there were about 50 clinical trials registered [4]. On February 25, 2020, the number of registered trials increased to 297. Before we finish the COS, we believe that it is very important to draw researchers’ attention to concern about outcomes in their research. So we conducted a review of outcome reporting from registered clinical trials of COVID-19. Because TCM and western medicine take the same role in the treatment of COVID-19 in China, so the review includes both interventions.

## METHODS

### Search strategy

All the databases of ICMJE-accepted clinical trial registry platform [6] were considered. Search terms for ChiCTR included “COVID-19”, “2019-novel Corona Virus (2019-nCoV)”, “Novel Coronavirus Pneumonia (NCP)”, “Severe Acute Respiratory Infection (SARI)”, “Severe Acute Respiratory Syndrome - Corona Virus-2 (SARS-CoV-2)”. Search terms for Netherlands National Trial Register (NTR) included “nCoV”, “Coronavirus”, “SARS”, “SARI”, “NCP”, “COVID”. Search terms for other databases included “2019-nCoV OR Novel Coronavirus OR New Coronavirus OR SARS-CoV-2 OR SARI OR NCP OR Novel Coronavirus Pneumonia OR COVID-19 OR Wuhan pneumonia”. The searching conducted on n February 14, 2020.

### Inclusion criteria

1. The population should include conformed patients of COVID-19.
2. Patients’ conditions include common type, severe type or critical type.
3. The interventions include any type of TCM therapy or western therapy.
4. The study types include randomized controlled trial (RCT) and observation study.

### Exclusion criteria

1. Studies for discharged patients.
2. Studies for psychological intervention.
3. Studies for complications of COVID-19.

### Study identification

Two reviewers (RQ and XW) independently assessed all the registered protocols. Any disagreement was resolved by discussion.

### Date extraction

Two reviewers (RQ and MZ) independently extracted information. The information included the primary investigators’ name, study type, type of disease, primary sponsor, number of settings, sample size, population’s age, course of treatment, interventions, outcomes, outcome definition/measurement instruments, measurement time frame. Any disagreement was resolved by discussion.

### Merging outcomes and grouping under outcome domains

Two researchers (RQ and CZ) merged the overlapping outcomes according to the definition of outcomes independently. If the researchers did not provide definition of outcome, they discussed and achieved consensus if necessary. For example, “PaO2/FiO2”, “oxygenation index”, “oxygen index”, “the difference of PaO2/FiO2 between two groups” were aggregated as “PaO2/FiO2”. Many protocols presented composite outcomes. If definitions were provided, or all of the single outcomes in the composite one can be measured in one test, it was listed in the review. If a single outcome which belongs to a composite outcome was reported by one or more protocols, the composite outcome was removed from the review. But when we conduct Delphi survey in further research, the composite outcome will be list to consult the participants’ opinion.

After the original outcomes were aggregated, two researchers (RQ and CZ) grouped individual outcomes into the appropriate outcome domain together and achieved consensus. The taxonomy of outcome domains were developed by the researchers from COMET initiative [7].

### Statistical analysis

The results were analysed by descriptive analysis.

### Patient and public involvement

The COVID-19 is highly infectious. For the safety of patients and public, they were not involved in the design or planning of the study.

## RESULTS

### Characteristics of literature

In this review, a total of 160 protocols from 19 different clinical trials registry platforms were searched. After reading titles and study details, 63 non-relevant or ineligible study protocols were excluded. In the end, 97 eligible study protocols were included from ChiCTR and ClinicalTrials.gov. The searching results and inclusion numbers were shown in Table 1.

**Table 1.**
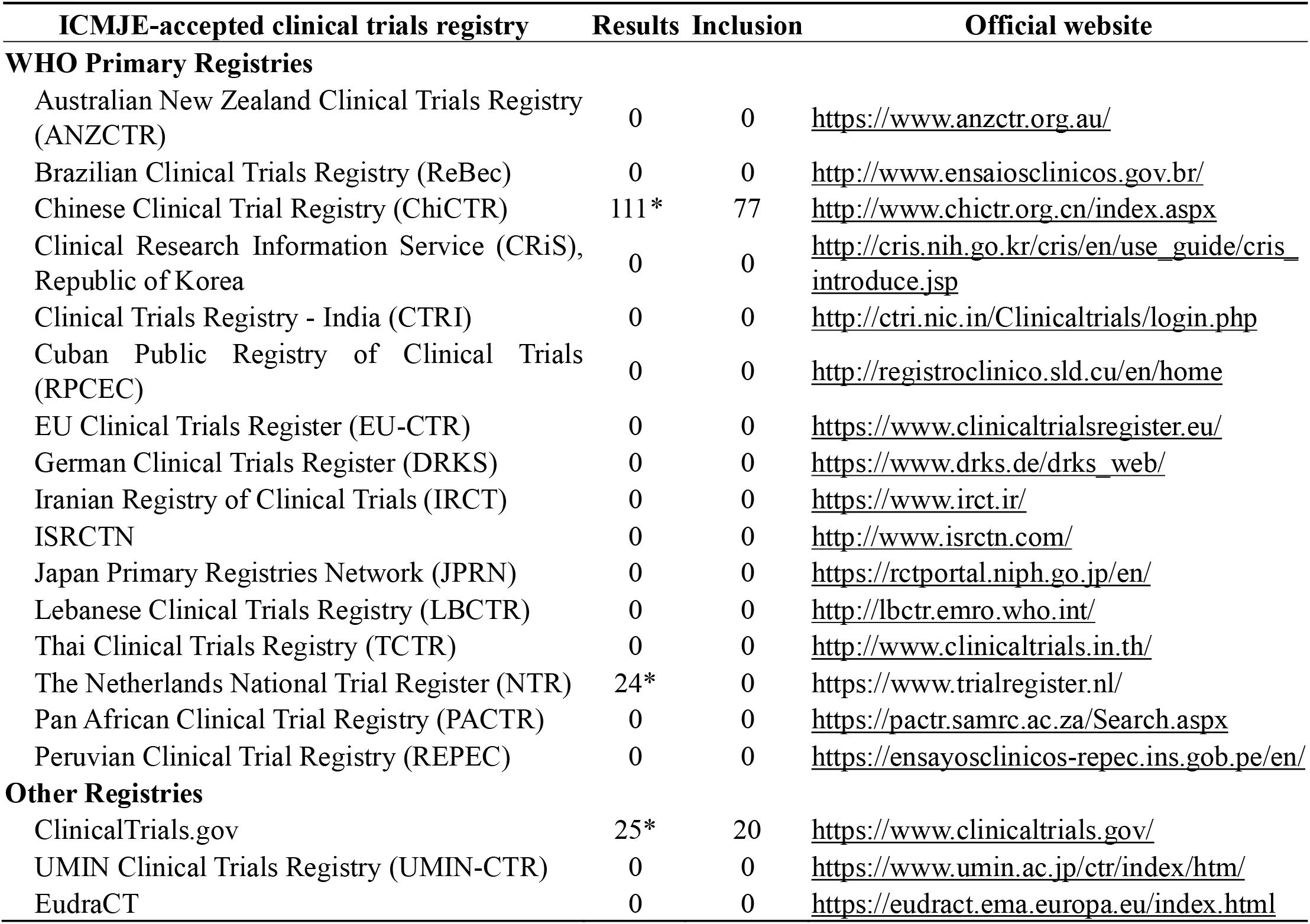
The global registry of COVID-19 related clinical trials searching results and inclusion.

In the included protocols, 34 clinical trials were for TCM therapy and 63 clinical trials were for western medicine therapy. All of clinical trials will be conducted in China. These clinical trials include 75 RCTs (53 for western medicine and 22 for TCM medicine) and 22 non-RCTs (10 for western medicine and 12 for TCM medicine). The first registered clinical trial for western medicine was on January 23, 2020, while the first registered clinical trial for TCM medicine was on January 27, 2020. The general characteristics of the included protocols are shown in table 2 and table 3.

**Table 2.**
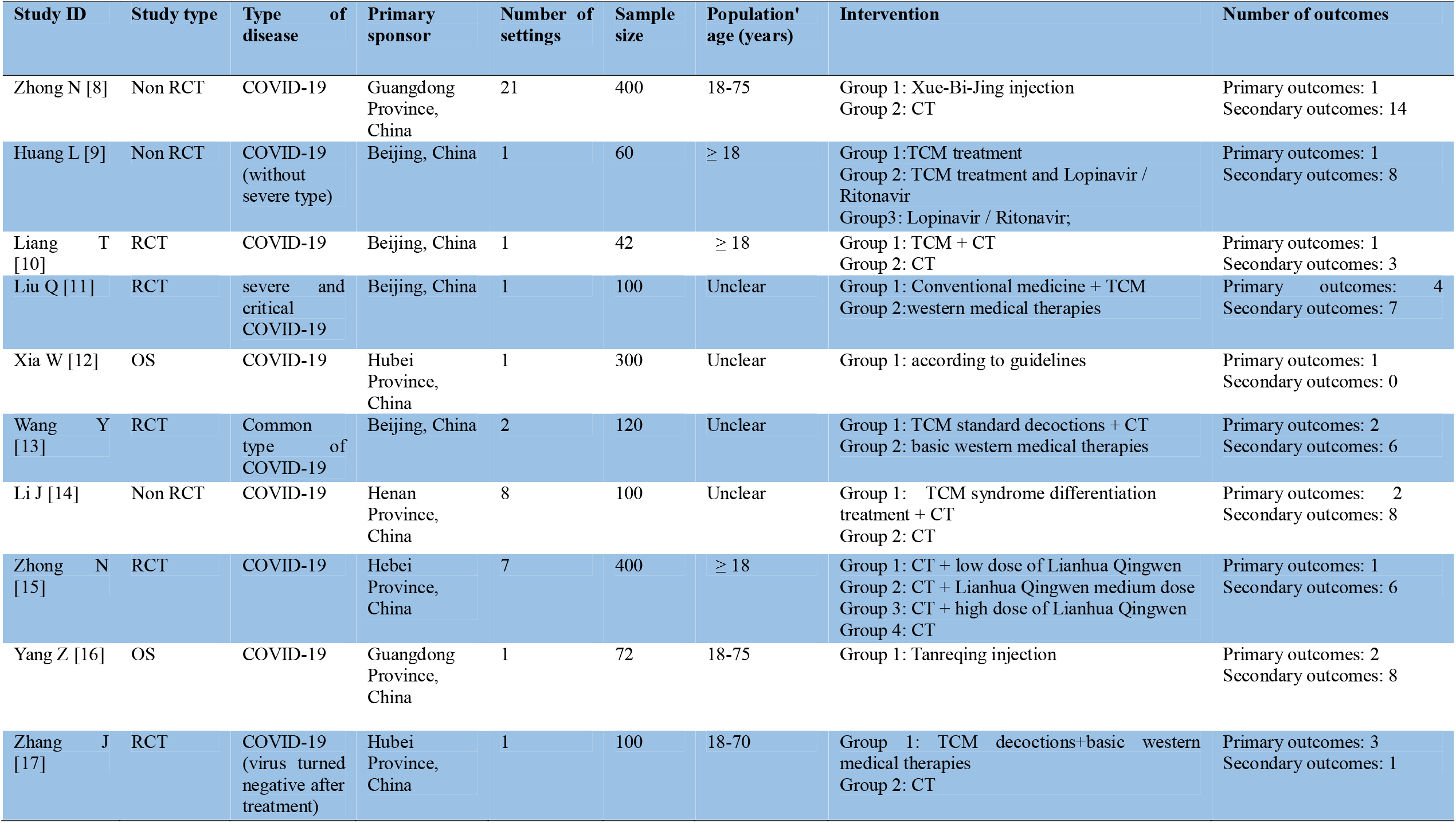
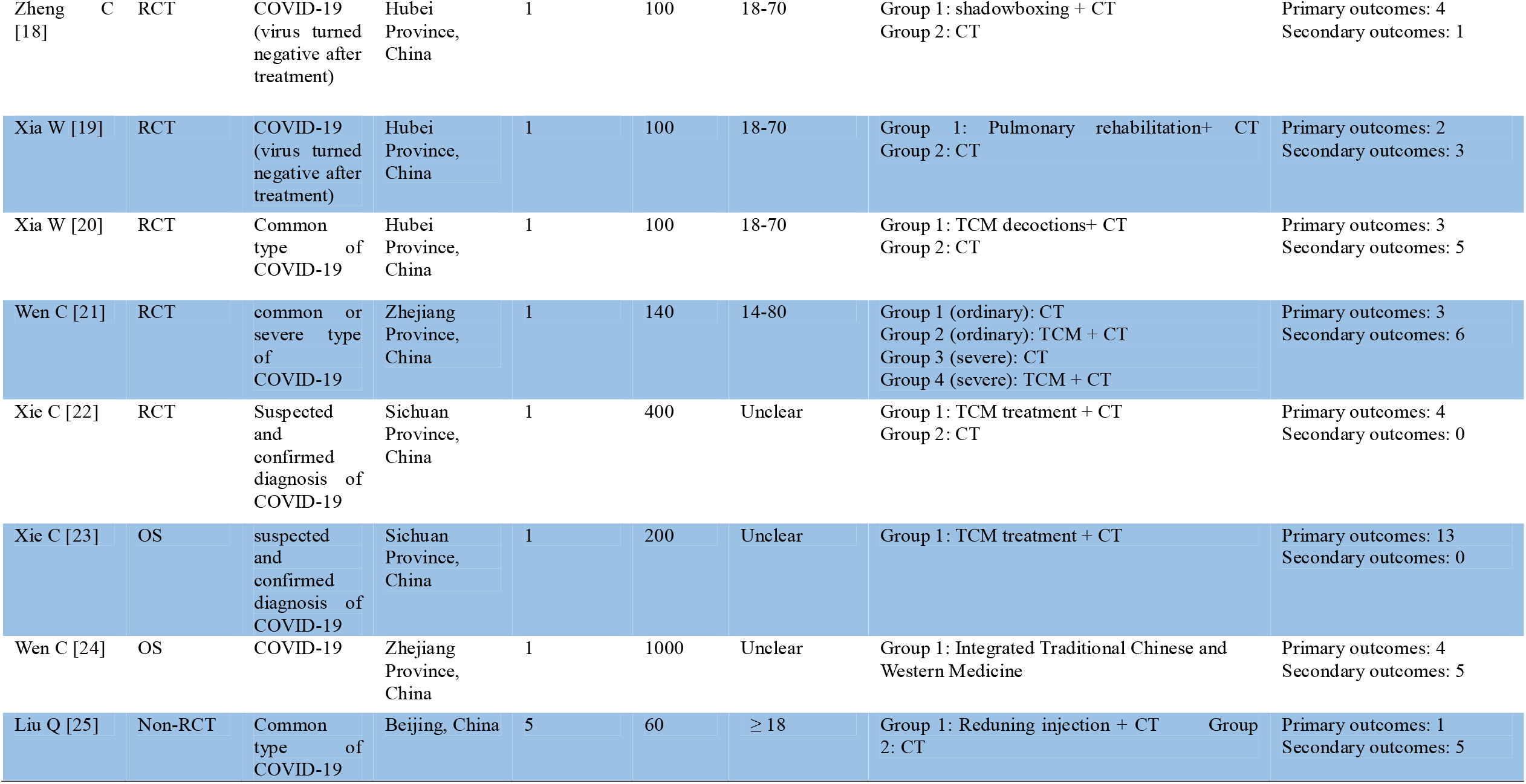
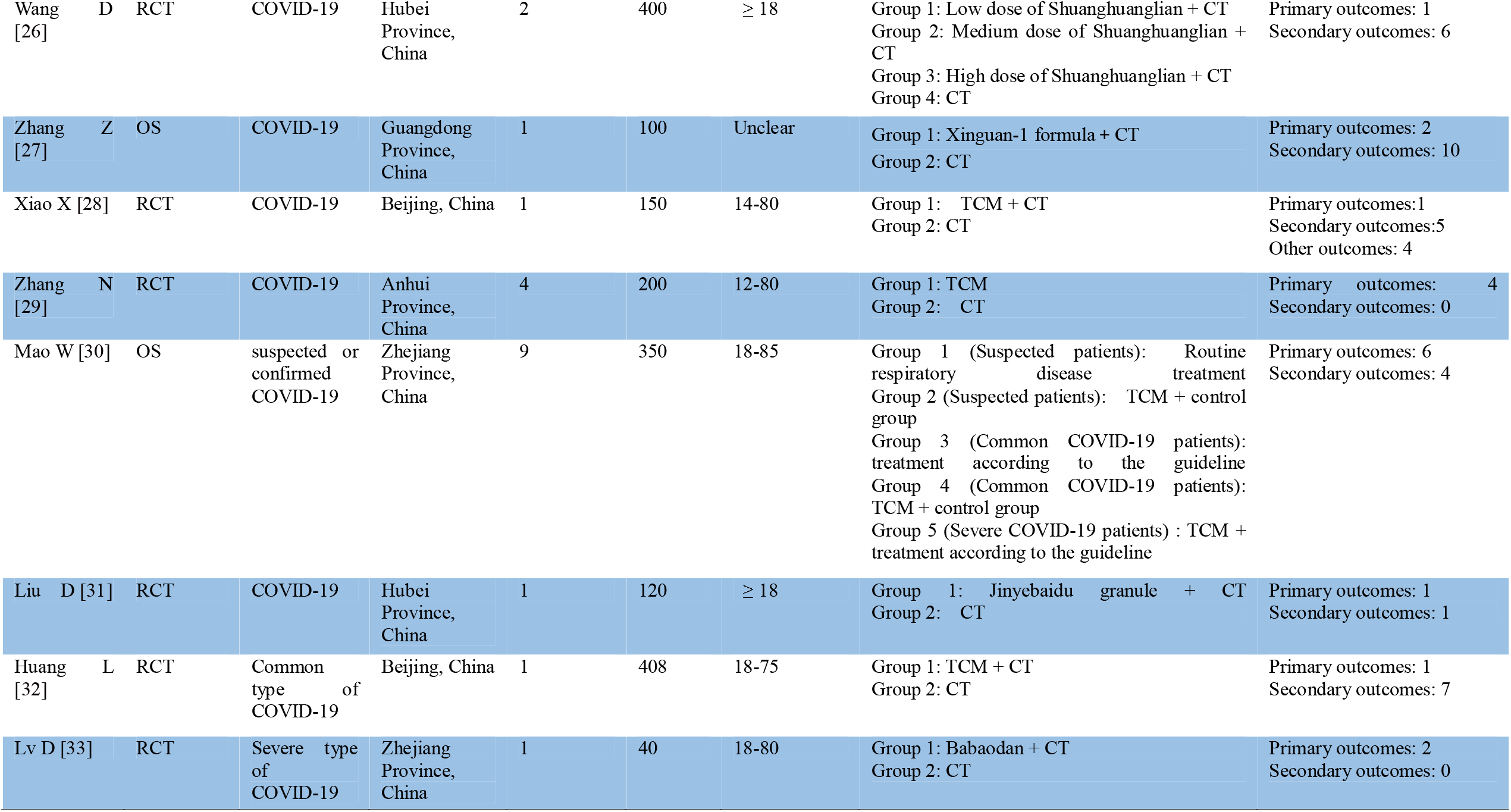
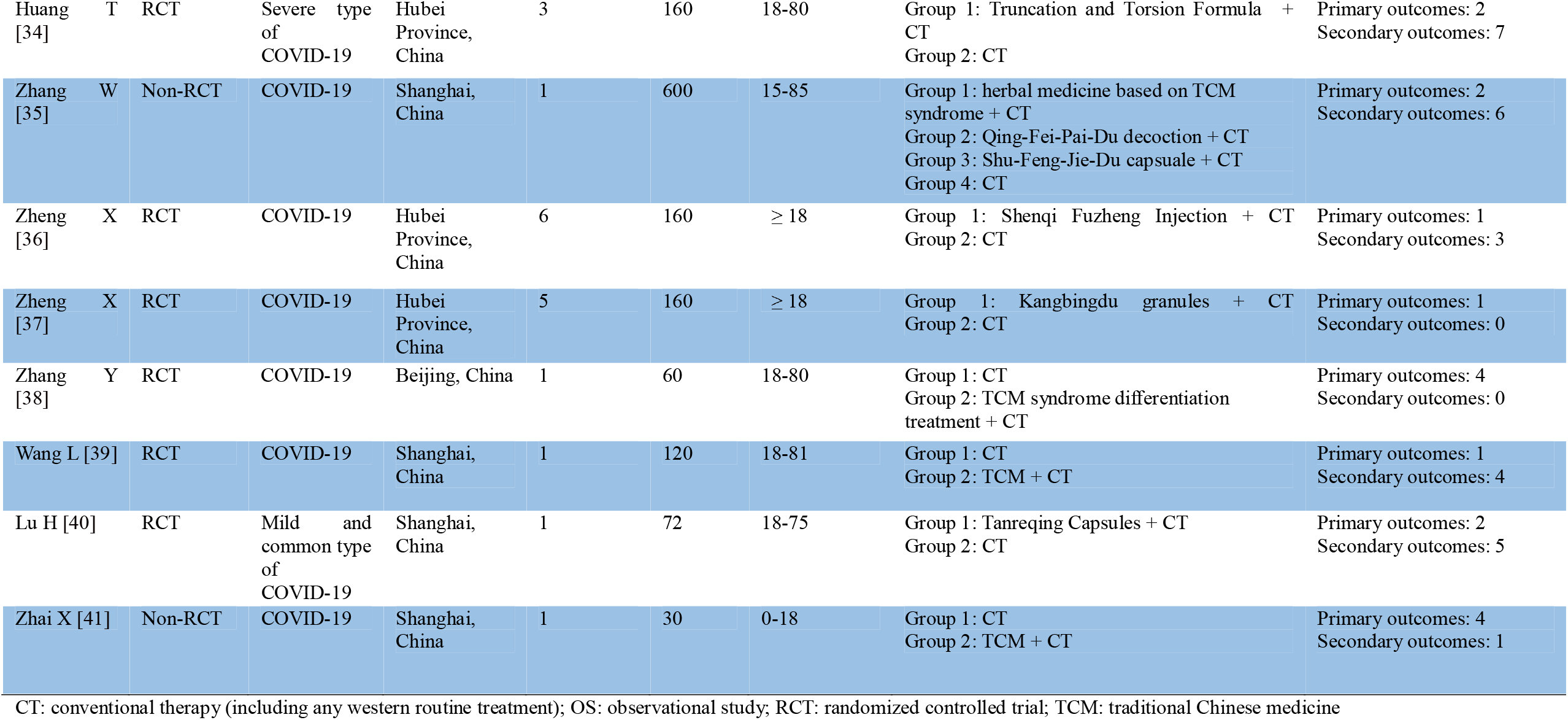
The characteristics of included protocols for TCM clinical trials.

**Table 3.**
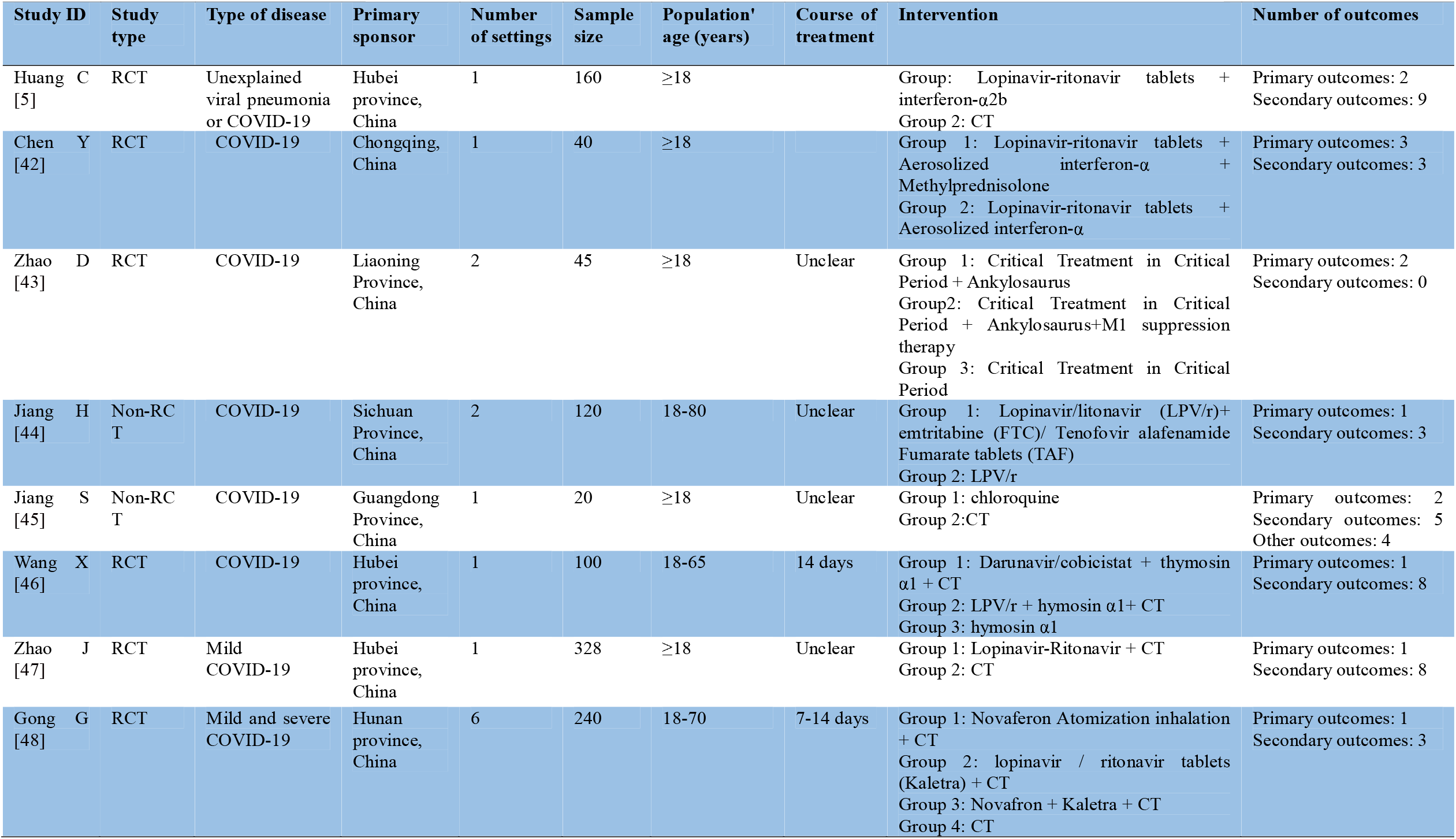
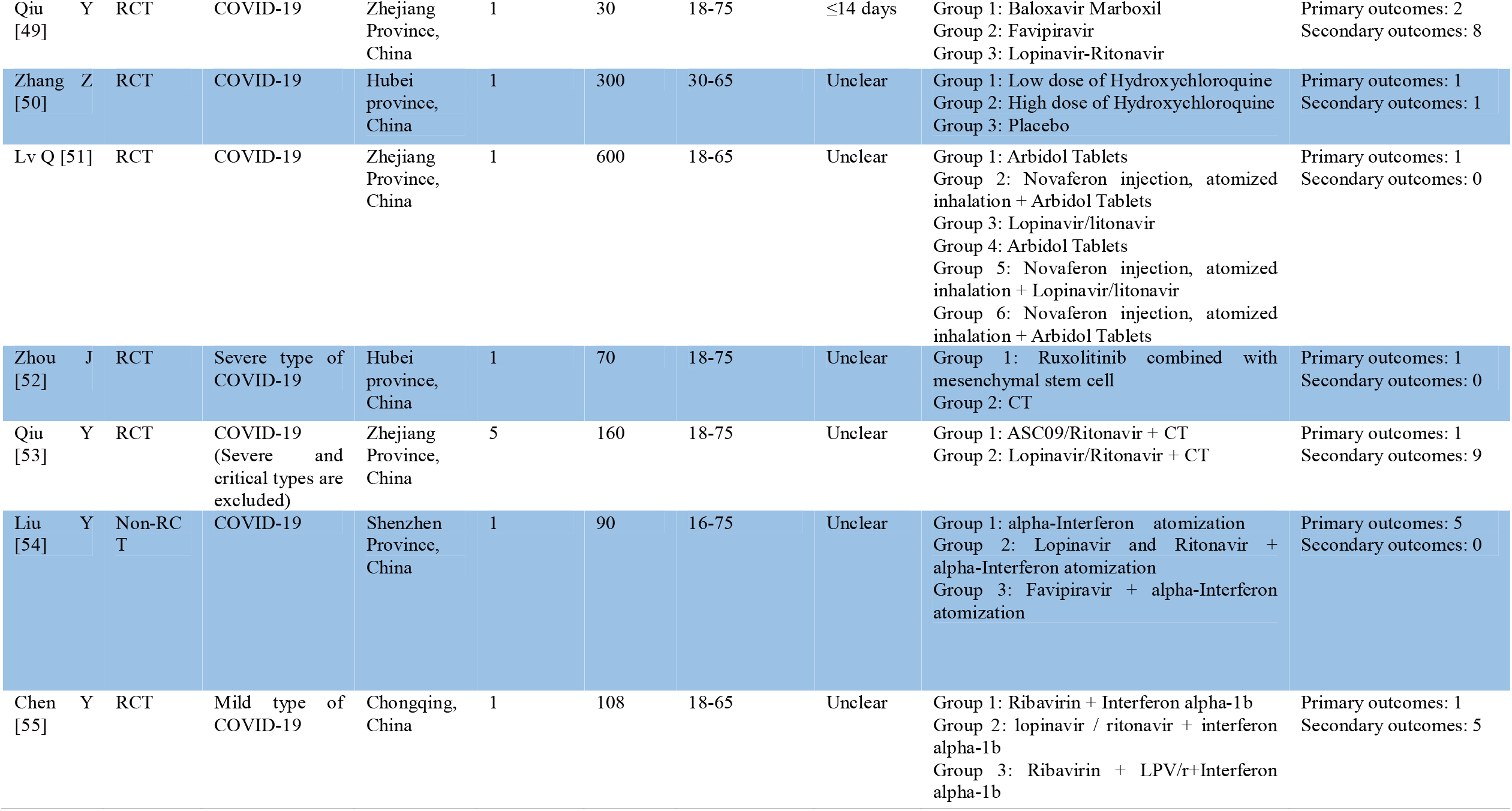
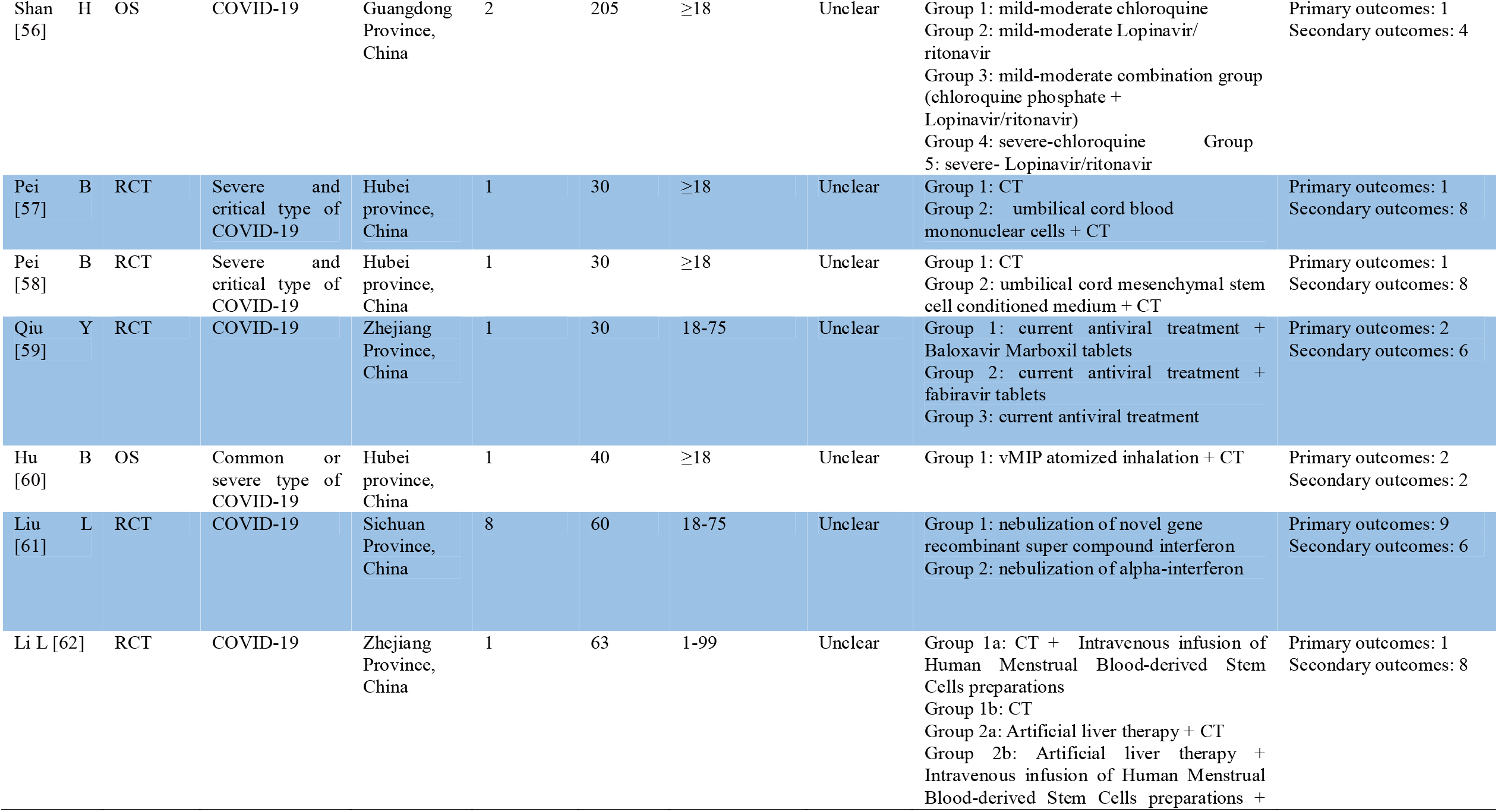
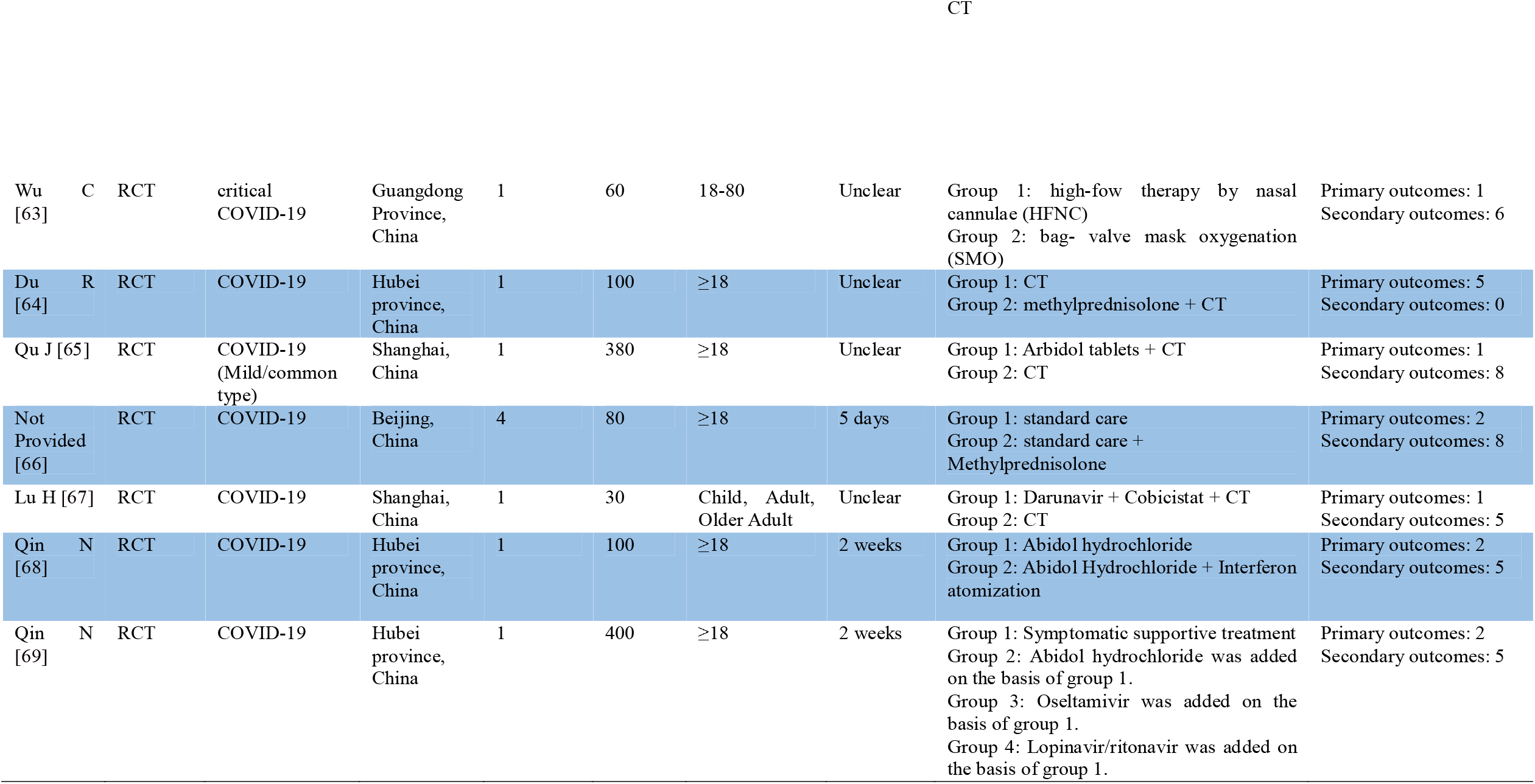
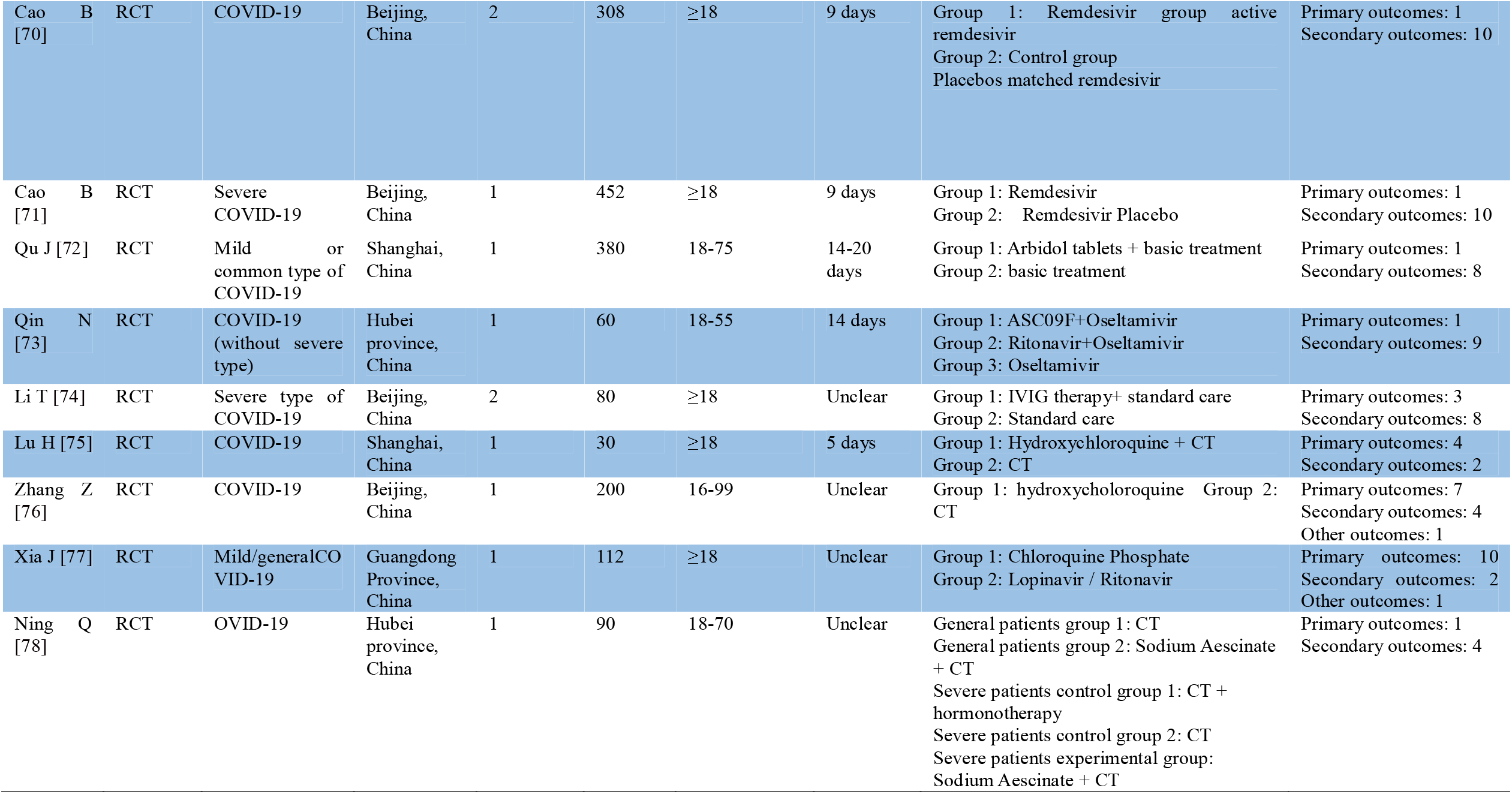
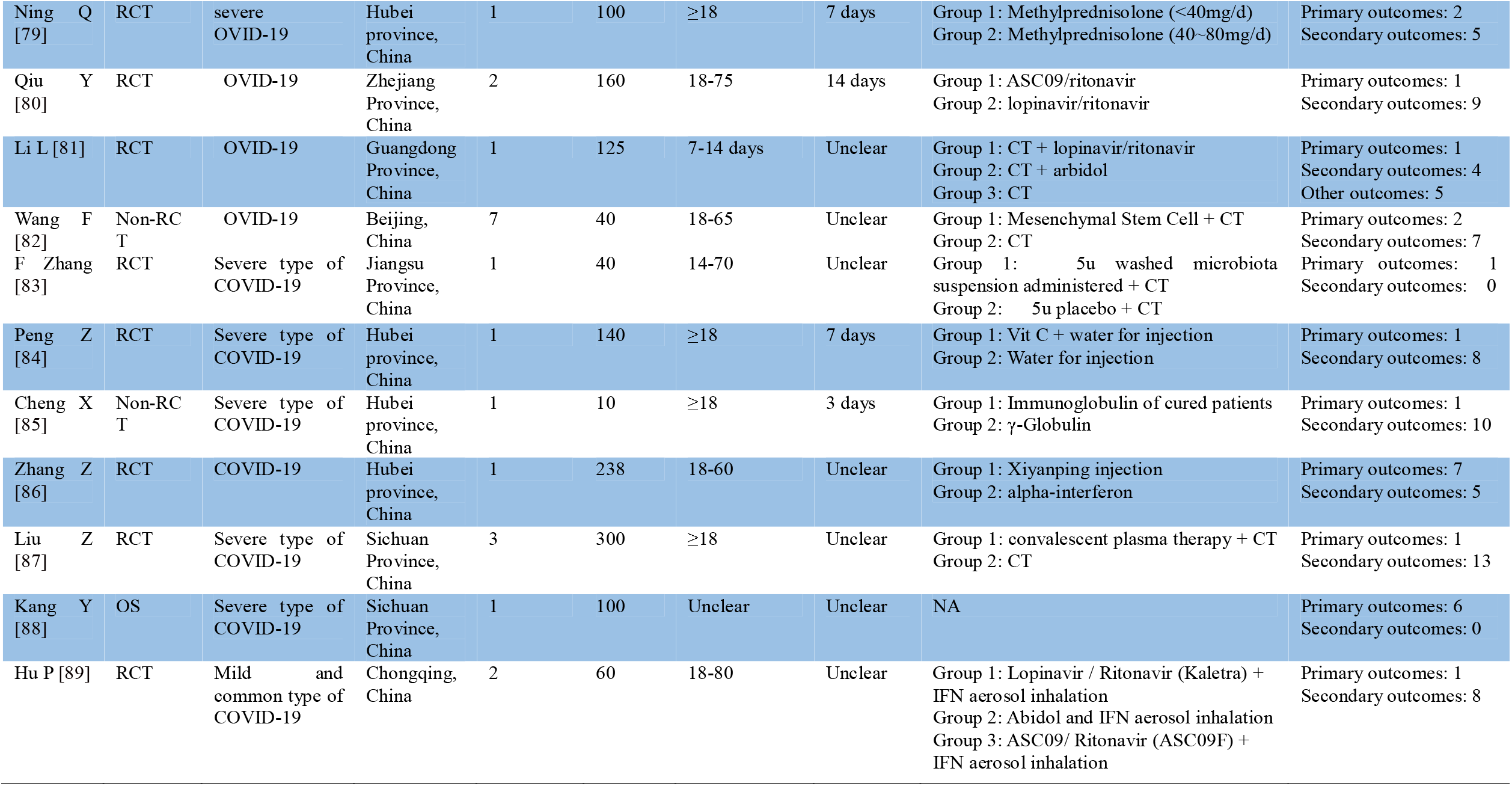
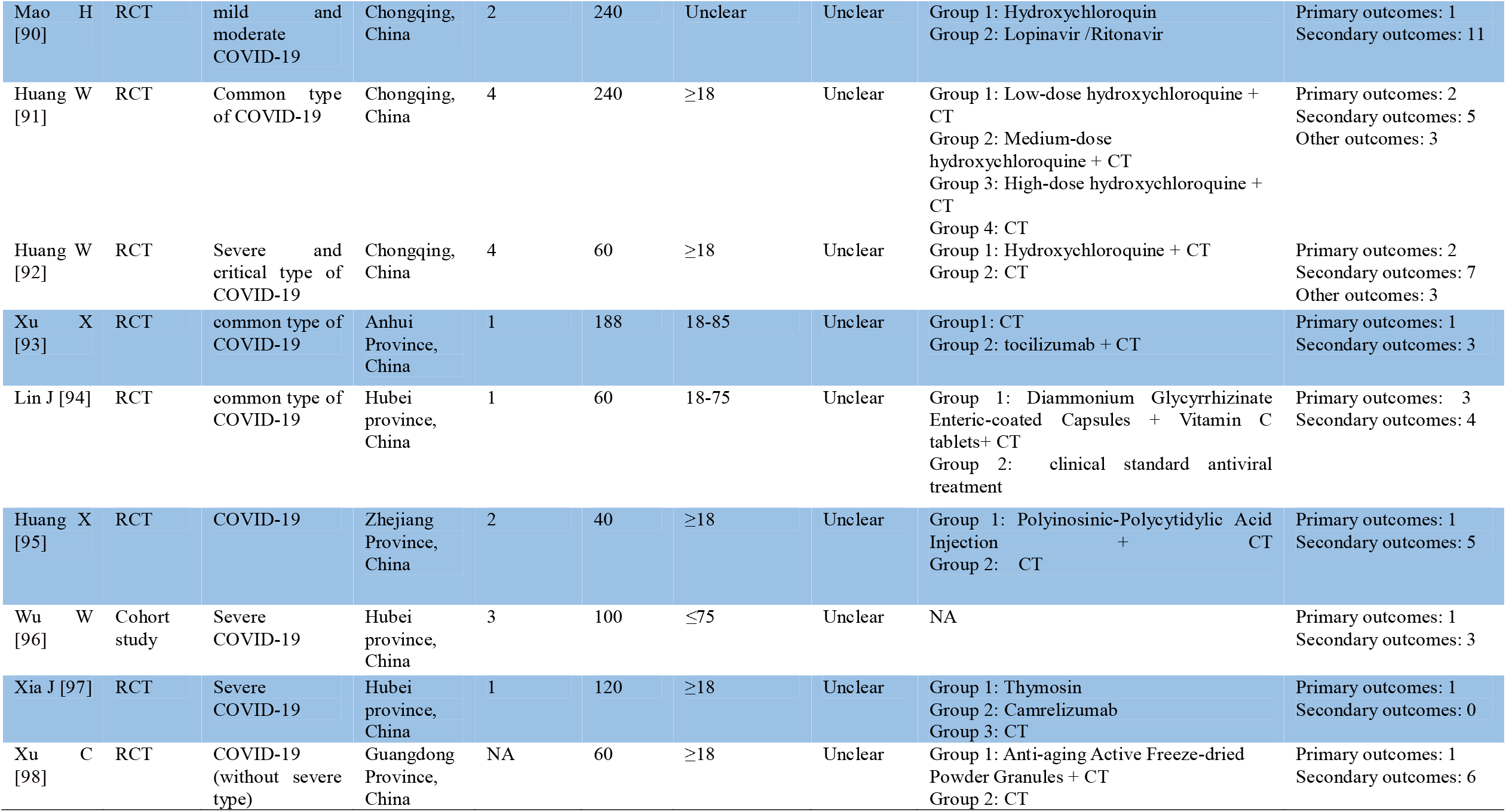
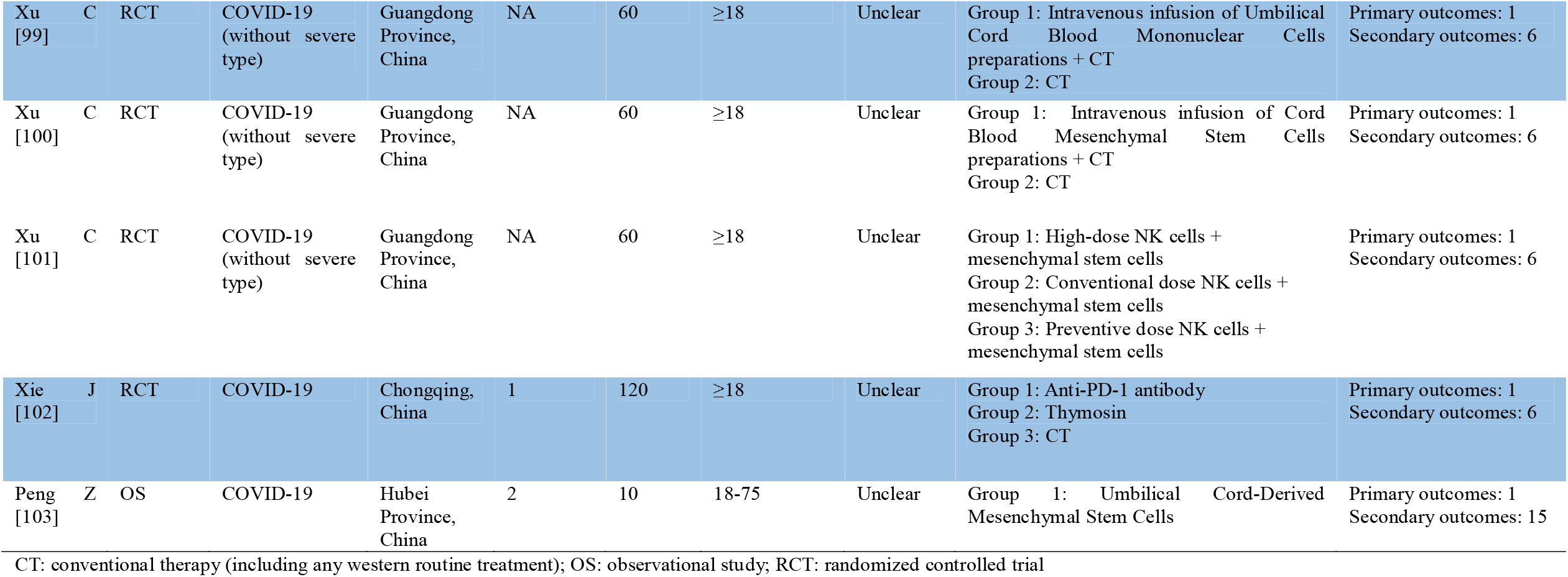
The characteristics of included protocols for western medicine clinical trials.

According to the information of primary sponsor, we found that the clinical trials were registered from 13 different provinces of China. Researchers from Hubei province registered more clinical trials (31/97, 31.96%) than researchers from other provinces. The distribution of clinical trials is shown in Figure 1.

**Figure 1.**
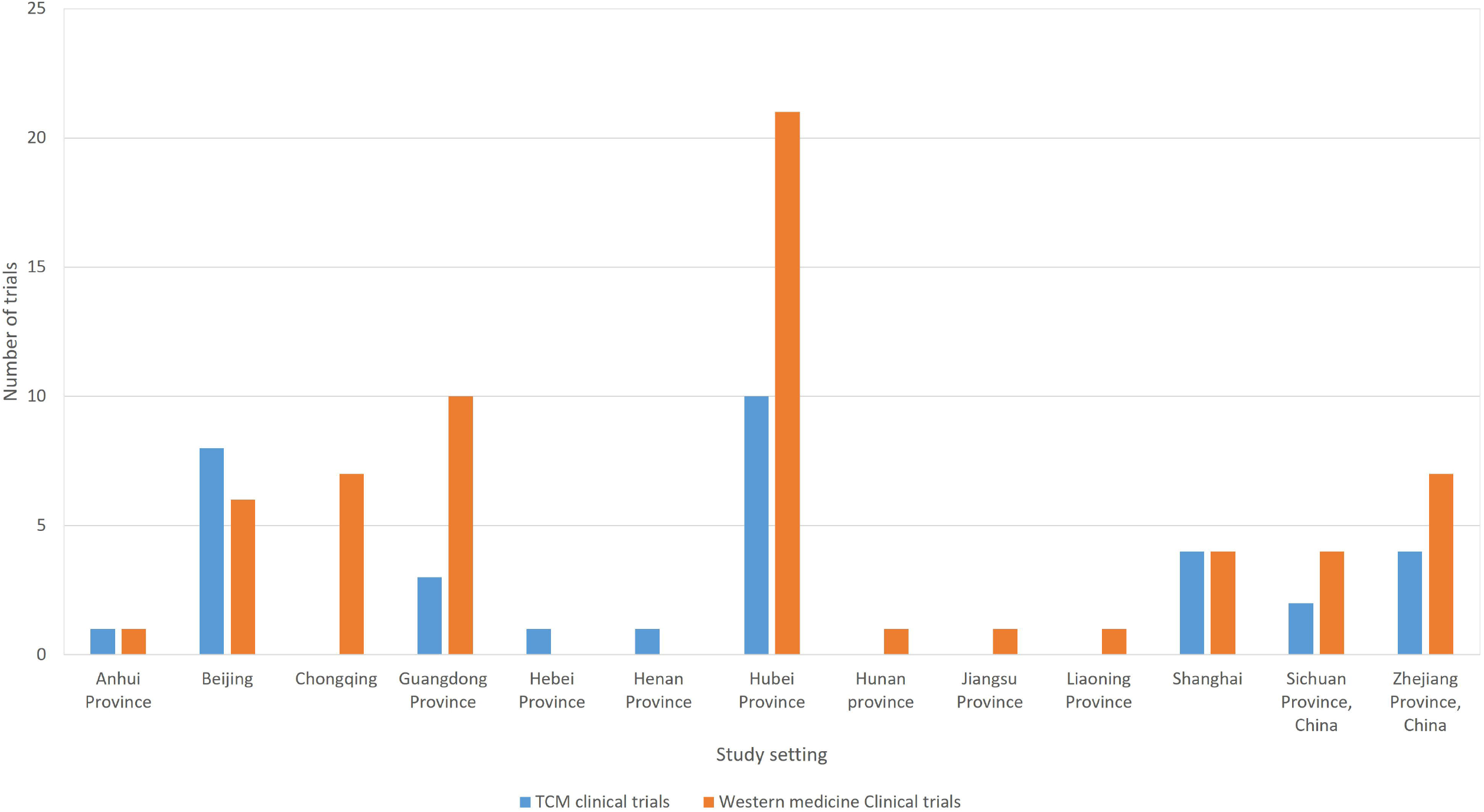
The distribution of clinical trials.

### The list of outcomes

In protocols of TCM clinical trials, the number of primary outcomes are from 1 (13/34, 38.24%) to 13 (1/34, 2.94%), the number of secondary outcomes are from 1 (7/34, 20.59%) to 14 (1/34, 2.94%). 1(1/34, 2.94%) protocol of clinical trial reports other outcomes. For individual clinical trial, the number of outcomes are from 1 (2/34, 5.88%) to 15 (1/34, 2.94%). The number of outcomes in protocols of TCM clinical trials is shown in Figure 2.

**Figure 2.**
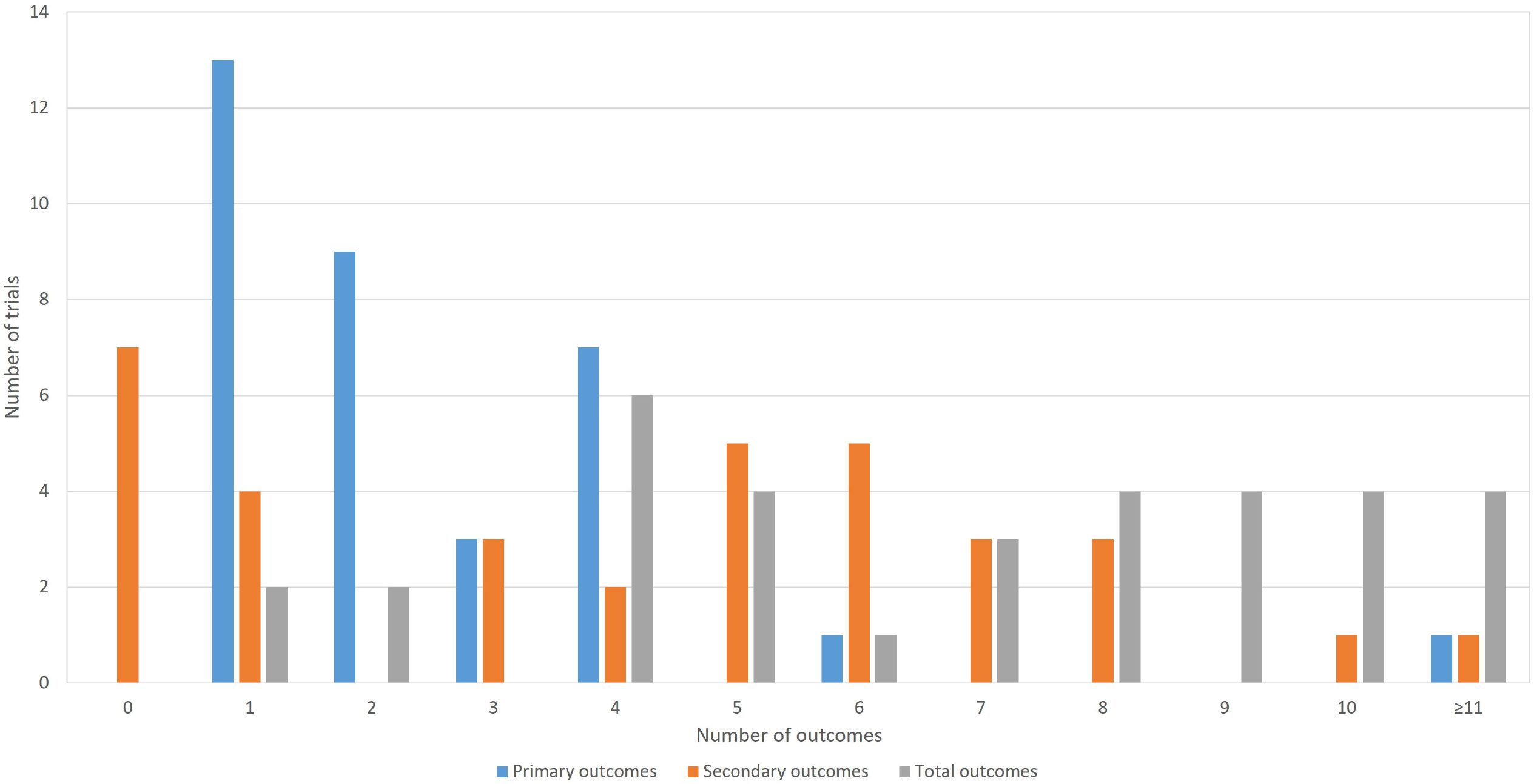
The number of outcomes in protocols of TCM clinical trials.

In protocols of western medicine clinical trials, the number of primary outcomes are from 1 (39/63, 61.90%) to 10 (1/63, 1.59%), the number of secondary outcomes are from 0 (8/63, 12.70%) to 15(1/63, 1.59%). 5 (5/63, 7.94%) protocols of clinical trials reported other outcomes (the number of outcomes are from 1 to 5). For individual clinical trial, the number of outcomes are from 1 (4/63, 6.35%) to 16 (1/63, 1.59%). The number of outcomes in protocols of western medicine clinical trials is shown in Figure 3.

**Figure 3.**
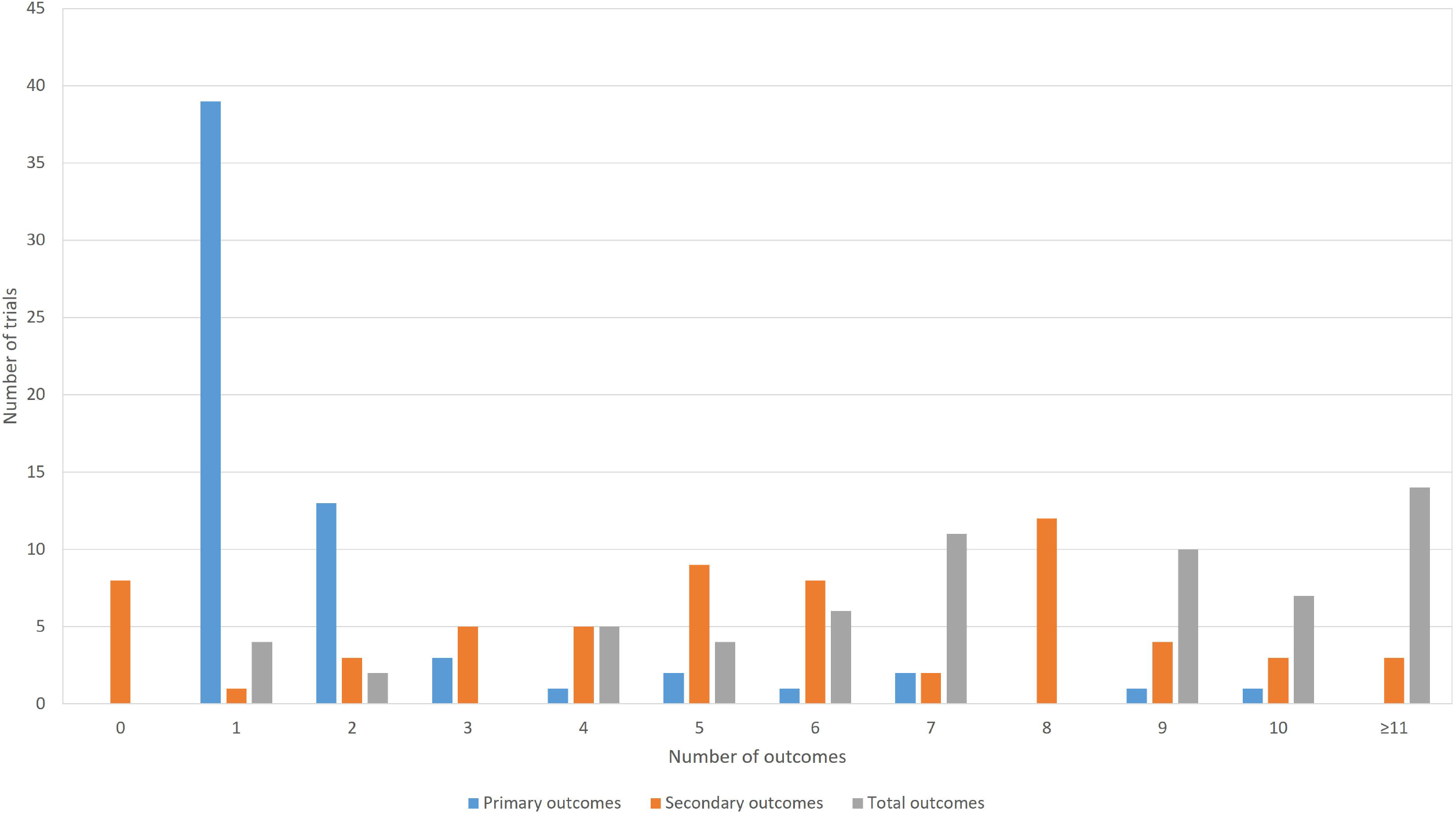
The number of outcomes in protocols of western medicine clinical trials.

After merging and grouping outcomes, there are 76 different outcomes from 16 outcome domains in 34 protocols of TCM clinical trials (table 4). Almost half of outcomes are reported only once (34/76, 44.74%). The most frequently reported outcome is “time of SARS-CoV-2 RNA turns to negative”, which is reported 16 times. Only 3 (3/76, 3.95%) outcomes are reported more than 10 times. Only 27 (27/76, 35.53%) outcomes are provided one or more outcome measurement instruments. Only 10 outcomes are provided one or more measurement time frame. The summary of outcome reporting for protocols of TCM clinical trials is shown in Figure 4.

**Table 4.** Outcomes from protocols of TCM clinical trials.

| Outcome domain | Outcomes | Number of outcomes | Defination/ outcome measurement | Time point |
| --- | --- | --- | --- | --- |
| **Mortality/survival** |  |  |  |  |
|  | Mortality/survival | 8 | 0 | 28 days, 84 days |
| **Physiological/clinical** |  |  |  |  |
| Blood and lymphatic system outcomes |  |  |  |  |
|  | Blood routine test | 10 | 0 | Not provided |
|  | Biochemical outcomes | 6 | 0 | Not provided |
|  | CRP | 5 | 0 | Not provided |
|  | Coagulation outcomes | 3 | 0 | Not provided |
|  | Erythrocyte sedimentation rate | 1 | 0 | Not provided |
|  | High-sensitive CRP | 1 | 0 | Not provided |
| **Cardiac outcomes** |  |  |  |  |
|  | ECG | 1 | 1 | Not provided |
|  | Heart function | 1 | 0 | Not provided |
|  | Heart rate | 1 | 0 | Not provided |
|  | Myocardial enzyme | 2 | 0 | Not provided |
|  | Myoglobin | 1 | 0 | Not provided |
|  | Troponin | 1 | 0 | Not provided |
| Gastrointestinal outcomes |  |  |  |  |
|  | Clearance time of gastrointestinal symptoms | 1 | 0 | Not provided |
|  | Remission of clinical symptoms: gastrointestinal discomfort | 4 | 0 | Not provided |
| General outcomes |  |  |  |  |
|  | Blood pressure | 1 | 0 | Not provided |
|  | Clearance time of fatigue | 1 | 0 | Not provided |
|  | Clearance time of fever | 14 | 1 | 1. 8 o'clock, 12 o'clock, 16 o'clock, and 20 o'clock, 2.28 days |
|  | Clinical symptom score | 1 | 0 | Not provided |
|  | Propotion of patients without fatigue | 9 | 0 | Not provided |
|  | Proportion of patients without fever | 3 | 0 | Not provided |
|  | Remission of clinical symptoms: fever | 4 | 0 | Not provided |
|  | Temperature | 3 | 0 | Not provided |
|  | TCM syndromes | 7 | 0 | Not provided |
|  | The proportion of patients without fever | 1 | 0 | Not provided |
|  | Time to remission/disappearance of primary symptoms | 1 | 1 | Not provided |
| Hepatobiliary outcomes |  |  |  |  |
|  | Liver function | 5 | 0 | Not provided |
| Immune system outcomes |  |  |  |  |
|  | HLA-DR | 1 | 0 | Not provided |
|  | Immunoglobulin | 1 | 0 | Not provided |
|  | Procalcitonin | 5 | 0 | Not provided |
|  | Rate of subjects receiving systematic corticosteroids | 1 | 1 | 28 days |
| Infection and infestation outcomes | Incidence of antibiotic treatment | 1 | 0 | Not provided |
|  | Proportion of patients with negative SARS-CoV-2 | 6 | 0 | at the end of the treatment, on the 28th day of treatment |
|  | Time of SARS-CoV-2 RNA turns to negative | 16 | 2 | Not provided |
| Renal and urinary outcomes | Urine routine | 2 | 0 | Not provided |
|  | kidney function | 5 | 0 | Not provided |
| Psychiatric outcomes |  |  |  |  |
|  | Psychological outcomes | 1 | 0 | Not provided |
| Respiratory, thoracic and mediastinal outcomes |  |  |  |  |
|  | PaO2/FiO2 | 4 | 0 | Not provided |
|  | Chest imaging | 12 | 1 | Not provided |
|  | Blood oxygen saturation | 3 | 0 | Not provided |
|  | CURB-65 | 1 | 1 | Not provided |
|  | Clearance time of cough | 4 | 0 | Not provided |
|  | Pulmonary function | 6 | 0 | Not provided |
|  | Duration of mechanical ventilation | 3 | 1 | 28 days |
|  | Improvement of lung HRCT score | 2 | 1 | Not provided |
|  | Pneumonia severity index | 4 | 1 | Not provided |
|  | Proportion of patients without cough | 6 | 0 | Not provided |
|  | Proportion of patients without sputum | 2 | 0 | Not provided |
|  | Proportion of patients without wheezing | 2 | 0 | Not provided |
|  | Respiratory rate | 1 | 0 | Not provided |
|  | St Georges respiratory questionnaire (SGRQ) | 4 | 1 | Not provided |
|  | Time to improvement of abnormalities in Chest radiology | 1 | 1 | 28 days |
|  | The incidence of dyspnea with low oxygen saturation level and high respiratory rate | 1 | 1 | 28 days |
|  | Time of improvement in respiratory symptoms | 1 | 0 | Not provided |
| **Functioning** |  |  |  |  |
| Physical functioning |  |  |  |  |
|  | 6-minute walk test (6MWT) | 4 | 1 | Not provided |
|  | APACHE II scores | 1 | 1 | Not provided |
|  | Clinical outcome | 1 | 1 | 14 day |
|  | DIC | 1 | 0 | Not provided |
|  | Modified Barthel Index (MBI) | 4 | 1 | Not provided |
|  | Major organ function | 1 | 0 | Not provided |
|  | Incidence of medical complications during hospitalization | 1 | 1 | Not provided |
|  | Incidence of multiple organ dysfunction | 1 | 0 | Not provided |
|  | Patients with complications of 2019-nCoV infection | 1 | 1 | 28 days |
|  | Recovery rate | 2 | 0 | Not provided |
|  | SOFA score | 1 | 1 | Not provided |
|  | Time to disease recovery | 5 | 0 | Not provided |
|  | Time to release from isolation | 2 | 0 | Not provided |
|  | Time to Clinical Recovery | 1 | 1 | Not provided |
| Global quality of life |  |  |  |  |
|  | EQ-5D | 1 | 1 | Not provided |
|  | SF-36 | 2 | 1 | Not provided |
| Resource use |  |  |  |  |
| Hospital |  |  |  |  |
|  | Duration of hospitalization | 10 | 1 | 28 days |
|  | Length of stay in ICU | 1 | 0 | Not provided |
| Need for further intervention |  |  |  |  |
|  | The time when the condition becomes worse | 6 | 3 | Not provided |
|  | Rate of progressing to the severe stage | 7 | 1 | Not provided |
|  | Rate of progressing to the critical stage | 10 | 1 | Not provided |
| Adverse events/effects |  |  |  |  |
|  | Adverse events | 9 | 0 | Not provided |

CRP: C-reactive protein; CURB-65: Confusion, Urea, Respiratory Rate and Age 65; ECG: electrocardiogram; HLA-DR: Human leukocyte antigen-DR; HRCT: chest high-resolution computed tomography; ICU: Intensive Care Unit; RNA: ribonucleic acid; SARS-CoV-2: Severe Acute Respiratory Syndrome Coronavirus 2; SOFA: Sequential Organ Failure Assessment; TCM: traditional Chinese medicine.

**Figure 4.**
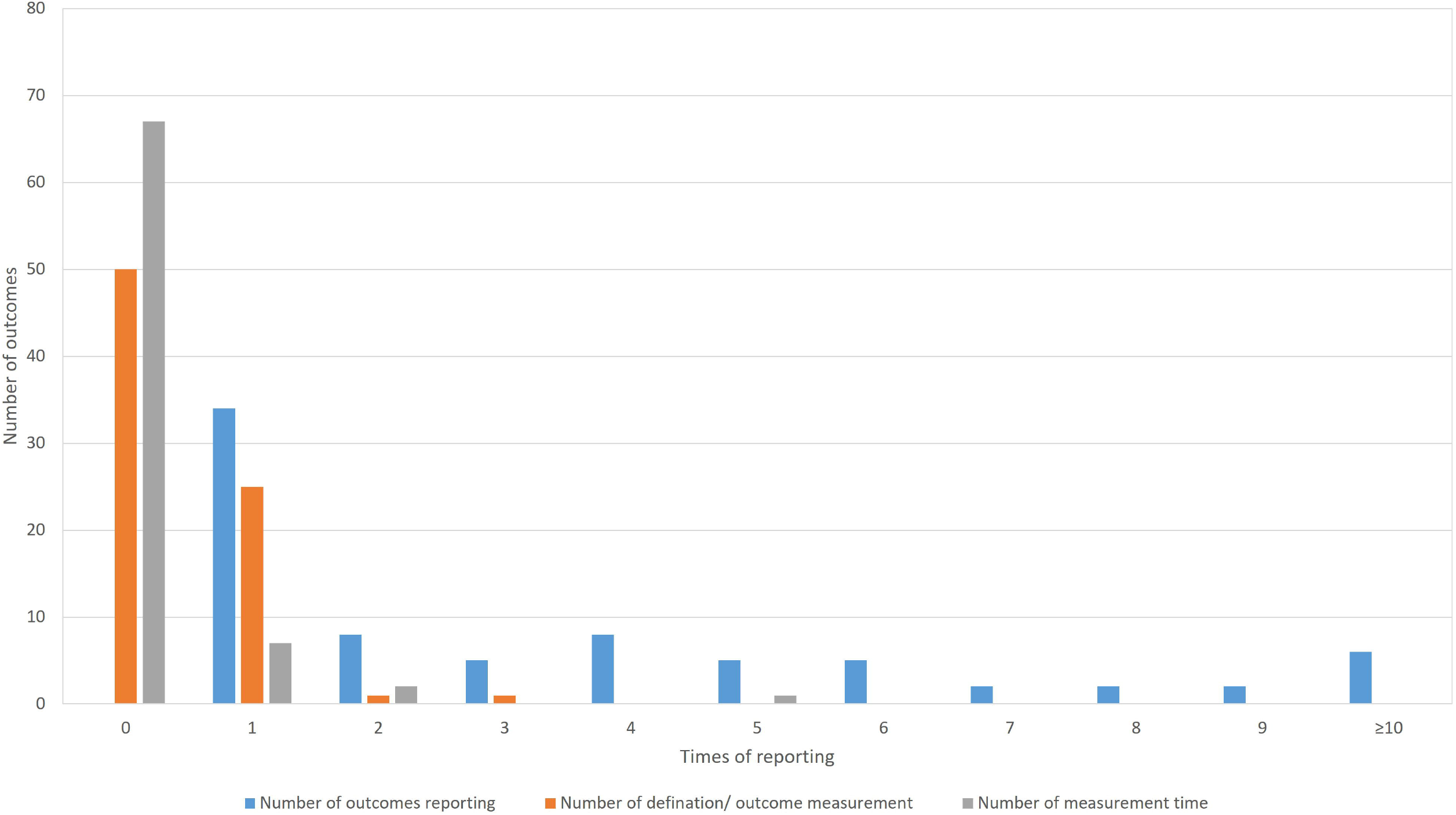
The summary of outcome reporting for protocols of TCM clinical trials.

In the 16 outcome domains of protocols of TCM clinical trials, 4 outcome domains (adverse events/effects, hepatobiliary outcomes, mortality/survival, psychiatric outcomes) consisted of only one outcome. These outcomes are reported between 1 and 9 times, and the median outcome reporting time was 6.5. Respiratory, thoracic and mediastinal outcomes have the largest number of outcomes, which includes 17 outcomes; chest imaging is reported more frequently than other outcomes. The number of outcomes in different outcome domains in protocols of TCM clinical trials is shown in Figure 5.

**Figure 5.**
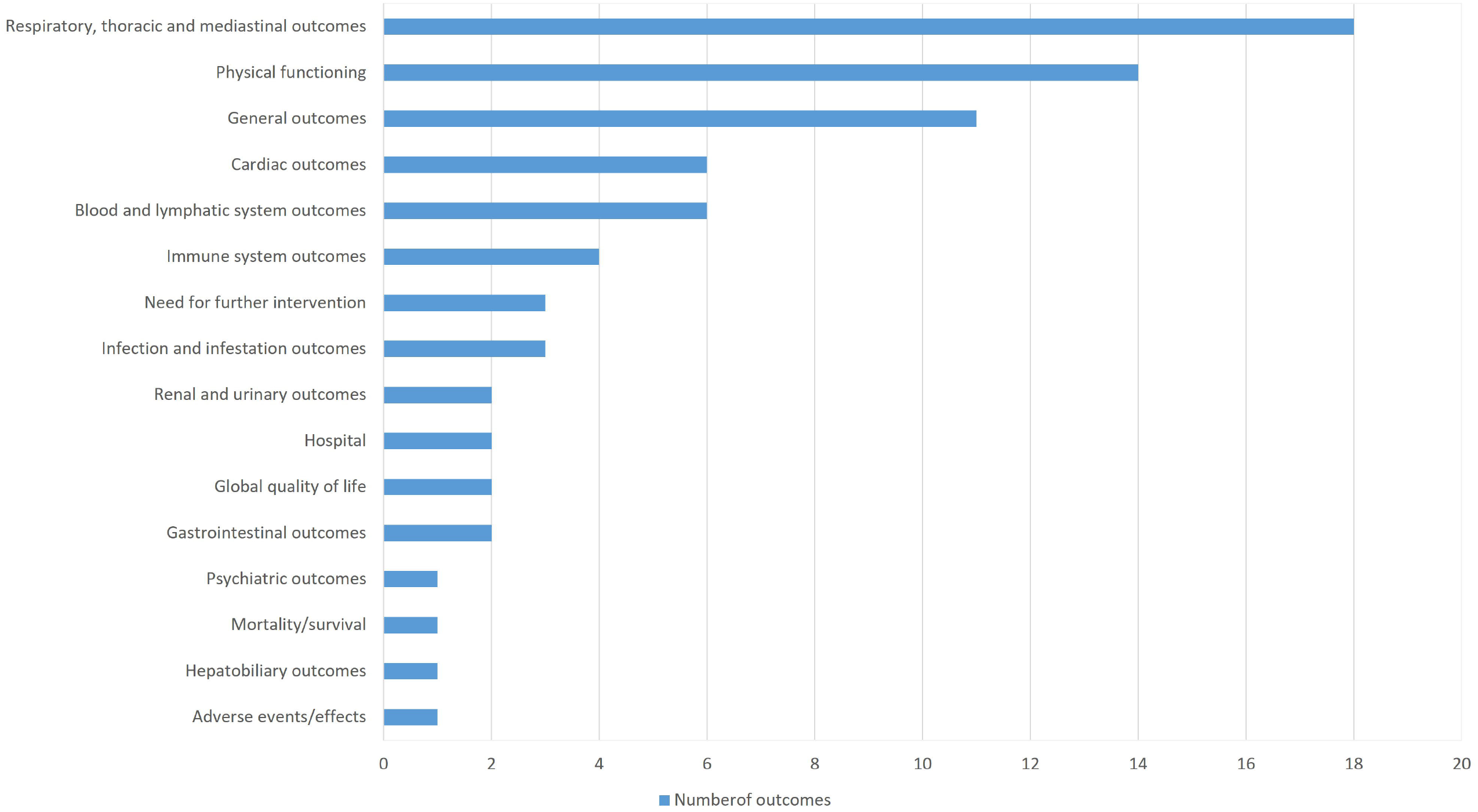
The number of outcomes in different outcome domains in protocols of TCM clinical trials.

After merging and grouping, there are 126 different outcomes from 17 outcome domains in 63 protocols of western medicine clinical trials (table 5). Almost half of outcomes are reported only once (62/126, 49.21%). The most frequently reported outcome is “proportion of patients with negative SARS-CoV-2”, which is reported 40 times. Only 11 (11/126, 8.73%) outcomes are reported more than 10 times. Only 27 outcomes are provided one or more outcome measurement instruments. Only 40 (40/126, 31.75%) outcomes are provided one or more measurement time frame. The summary of outcome reporting for protocols of TCM clinical trials is shown in Figure 6.

**Table 5.** Outcomes from protocols of western medicine clinical trials.

| Outcome domain | Outcomes | Number of outcomes | Definition/ outcome measurement | Time point |
| --- | --- | --- | --- | --- |
| **Mortality/survival** |  |  |  |  |
|  | Mortality/survival | 36 | 1 | 2 weeks, 4 weeks, 12 weeks, 14-20 days, 28 days. |
| **Physiological/clinical** |  |  |  |  |
| Blood and lymphatic system outcomes |  |  |  |  |
|  | Blood routine test | 16 | 1 | At baseline, day 1, 3, 4, 5, 7, 10, 14, 21, 28. |
|  | Biochemical outcomes | 7 |  | Not provided |
|  | CRP | 9 | 0 | At Baseline, day1, 3, 5, 6, 7, 10,14, 21, 28, 90 |
|  | Time of CRP recovery | 1 | 0 | 28 days |
|  | Coagulation outcomes | 2 | 1 | Day 1, 3, 5, 7, 10, 14, 21, 28 |
|  | ES rate | 1 | 0 | Not provided |
|  | Rate of ES recovery | 4 | 0 | Two weeks, 28 days |
|  | Time of ES recovery | 1 | 0 | 28 days |
| **Cardiac outcomes** |  |  |  |  |
|  | ECG | 3 | 1 | Not provided |
|  | Heart rate | 6 | 0 | Baseline, week 1, week 2, week 3, week 4 |
|  | Myocardial enzymes | 2 | 0 | Day 0, 3, 4, 6, 7, 10, 14, 28, 90 |
|  | Mb | 1 | 0 | Day 1, 3, 5, 7, 10, 14, 21, 28 |
|  | Rate of Mb recovery | 4 | 0 | Two weeks, 28 days |
|  | Rate of CK recovery | 4 | 0 | Two weeks, 28 days |
|  | Time of Mb recovery | 1 | 0 | 28 days |
|  | Time of CK recovery | 1 | 0 | 28 days |
| General outcomes |  |  |  |  |
|  | Abnormal time of temperature during infection | 1 | 0 | Not provided |
|  | Antipyretic rate | 2 | 1 | 14-20 days |
|  | Blood pressure | 6 | 0 | Day 0 till day 21 |
|  | Clearance time of fever | 15 | 0 | 14-20 days, up to 28 days |
|  | Clearance time of fatigue | 6 | 0 | Not provided |
|  | Clearance time of myalgia | 2 | 0 | 14-20 days |
|  | Proportion of patients without fatigue | 6 | 0 | Not provided |
|  | Proportion of patients without dyspnea | 5 | 0 | 14 days |
|  | Proportion of patients without fever | 14 | 0 | Day 7, within 14 days |
|  | Temperature | 6 | 0 | Baseline, 1 week, 2 weeks, 3 weeks, 4weeks |
| Hepatobiliary outcomes |  |  |  |  |
|  | ALT | 1 | 0 | At baseline , day 3, 6, 10, 14, 28 and 90 |
|  | Rate of ALT recovery | 4 | 0 | Two weeks, 28 days |
|  | Time to ALT recovery | 1 | 0 | 28 days |
| Immune system outcomes |  |  |  |  |
|  | CD4+ T cell count | 1 | 0 | At Baseline , day 3, 6, 10, 14, 28 and 90 |
|  | CD8+ T cell count | 1 | 0 | At Baseline , day 3, 6, 10, 14, 28 and 90 |
|  | IL-2 | 1 | 0 | Day 7, 14, 28 |
|  | IL-4 | 1 | 0 | Day 7, 14, 28 |
|  | IL-6 | 8 | 0 | Day 1, 3, 5, 7, 10, 14, 21, 28 |
|  | IL-8 | 1 | 0 | On the Day28 |
|  | IL-10 | 1 | 0 | Day 7, 14, 28 |
|  | Immunoglobulin | 1 | 0 | Not provided |
|  | Lymphocyte subsets and complement | 1 | 0 | Day 1, 3, 5, 7, 10, 14, 21, 28 |
|  | Procalcitonin | 4 | 0 | Day 1, 3, 5, 7, 10, 14, 21, 28 |
|  | Recovery time of lymphocyte | 1 | 0 | Not provided |
|  | Time to CD4+ T cell recovery | 1 | 0 | Not provided |
|  | Time to CD8+ T cell recovery | 1 | 0 | Not provided |
| | TNF-α | 4 | 0 | Day 7, 14, 28 |
| | γ-interferon | 1 | 0 | Day 7,14, 28 |
| Infection and infestation outcomes |  |  |  |  |
|  | Chloroquine blood concentration | 1 | 0 | Day 1, 3, 5, 7, 10, 14, 21, 28 |
|  | Declining speed of SARS-CoV-2 | 1 | 1 | Not provided |
|  | Duration of antibiotic treatment | 1 | 0 | Not provided |
|  | Hemodiafiltration | 1 | 0 | Not provided |
|  | Incidence of antibiotic treatment | 2 | 0 | Not provided |
|  | Level of virus antibody in blood sample | 1 | 0 | Not provided |
|  | Other infection | 1 | 0 | Not provided |
|  | Proportion of patients with negative SARS-CoV-2 | 40 | 1 | Day 0, 1, 2, 3, 4, 5, 7, 10, 14, 16, 21, 28 |
|  | Time of SARS-CoV-2 RNA turns to negative | 23 | 1 | At baseline, day 3, 6, 7, 10, 14, 28, 90 |
| Metabolism and nutrition outcomes |  |  |  |  |
|  | Liquid balance | 1 | 0 | Not provided |
| Musculoskeletal and connective tissue outcomes |  |  |  |  |
|  | MRI of hip | 1 | 1 | Not provided |
|  | CT of hip | 1 | 1 | Not provided |
| Renal and urinary outcomes |  |  |  |  |
|  | Urine routine | 1 | 0 | Not provided |
|  | kidney function | 2 | 1 | Day 0, 1, 3, 4, 5, 7, 10, 14, 21, 28 |
|  | Incidence rate of kidney damage | 1 | 0 | Not provided |
| Respiratory, thoracic and mediastinal outcomes |  |  |  |  |
|  | Application of pulmonary surfactant | 1 | 0 | Not provided |
|  | Blood oxygen saturation | 2 | 0 | Day 0 till day 21 |
|  | PaO2/FiO2 | 7 | 1 | Day 1, 3, 5, 7, 10, 14, 21, 28 |
|  | Chest imaging | 25 | 2 | At baseline , day 3, 4, 6, 7, 10, 14, 21, 28 |
|  | Clearance time of cough | 9 | 0 | 14-20 days |
|  | Clearance time of dyspnea | 3 | 0 | 14-20 days, up to 28 days |
|  | Duration of extracorporeal membrane oxygenation | 2 | 0 | Up to 28 days |
|  | Duration of supplemental oxygenation | 6 | 0 | Up to 28 days |
|  | Duration of mechanical ventilation | 10 | 0 | From Day 0 through Day 28 |
|  | Finger oxygen improvement rate | 2 | 1 | 14-20 days |
|  | Frequency of requirement for mechanical ventilation | 1 | 0 | Up to 28 days |
|  | Frequency of requirement for supplemental oxygen | 1 | 0 | 14 days, 28 days |
|  | Frequency of respiratory progression | 2 | 1 | Up to 28 days |
|  | Murray lung injury score | 5 | 1 | 7 days, 14 days |
|  | Oxygen intake methods | 1 | 1 | Up to 28 days |
|  | Pneumonia severity index | 2 | 0 | Not provided |
|  | Proportion of patients without cough | 12 | 0 | Within 14 days, up to 28 days |
|  | Respiratory rate | 6 | 1 | Baseline, 1 week, 2 weeks, 3weeks, 4weeks |
|  | Rate of mechanical ventilation | 9 | 1 | Day 7, 14, 15 |
|  | Time to normalization of respiratory rate | 1 | 0 | Not provided |
|  | The duration of intubation | 1 | 0 | Not provided |
|  | The incidence of hypoxia | 1 | 0 | Not provided |
|  | The lowest SpO2 during intubation | 1 | 0 | Not provided |
|  | Time to chest imaging recovery | 2 | 1 | Two weeks |
|  | The times of intubation | 1 | 0 | Not provided |
|  | Time of improvement in respiratory symptoms | 2 | 0 | Not provided |
|  | Time of using assisted breathing | 2 | 0 | Not provided |
|  | Time to cough reported as mild | 1 | 0 | Not provided |
|  | Time to dyspnea reported as mild | 1 | 1 | Up to 28 days |
|  | Rate of no requiring supplemental oxygen | 11 | 0 | 14 days |
|  | Ventilator parameters | 5 | 1 | Day 10 and 28 |
| **Functioning** |  |  |  |  |
| Physical functioning |  |  |  |  |
|  | 7-point scale | 1 | 1 | Not provided |
|  | APACHE II scores | 1 | 1 | Day 10 |
|  | Complications | 2 | 0 | Not provided |
|  | Demand for first aid measurements | 1 | 1 | On the day 28 |
|  | Disease progression rate | 4 | 1 | 14-20 days |
|  | Incidence of multiple organ dysfunction | 1 | 0 | Not provided |
|  | Incidence of shock | 1 | 0 | Not provided |
|  | Lower SOFA score | 2 | 1 | 7 days, 14 days |
|  | NEWS2 score | 1 | 1 | Not provided |
|  | Number of participants with improvement from severe type to common type | 1 | 1 | 2 weeks |
|  | Organ function support measures | 1 | 0 | Not provided |
|  | Organ support intensity | 1 | 0 | Not provided |
|  | Rate of severe type of disease | 2 | 0 | Not provided |
|  | Rate of preventing mild to moderate patients from shifting to severe patients | 1 | 0 | Not provided |
|  | Rate of disease remission | 5 | 1 | Day 7, within 14 days |
|  | SOFA score | 3 | 1 | Day 7, 10 |
|  | Time to treatment failure | 2 | 1 | Not provided |
| | Time to NEWS2 of $\leq 2$ maintained for 24 hours. | 1 | 0 | Up to 28 days |
|  | Time to release from isolation | 1 | 0 | Not provided |
|  | Time to severe stage | 2 | 0 | Not provided |
|  | The rate of critical stage | 3 | 1 | 2 weeks |
|  | Time to Clinical Recovery | 7 | 4 | 14 days, 28 days |
|  | Time to Clinical Improvement | 8 | 2 | Up to 28 days |
| Delivery of care |  |  |  |  |
|  | The rate of discontinuations due to adverse events | 1 | 0 | Not provided |
| Resource use |  |  |  |  |
| Economic | Hospitalization costs | 1 | 0 | Not provided |
| Hospital |  |  |  |  |
|  | Duration of hospitalization | 23 | 0 | From day 0 through day 28 |
|  | Length of stay in ICU | 10 | 1 | Day 28 |
|  | ICU free days | 1 | 0 | Up to 28 days |
|  | Incidence of ICU admission | 9 | 0 | Day7, 14, 15, 28 |
|  | The proportion of inpatients | 1 | 0 | Within 28 days |
| Need for further intervention |  |  |  |  |
|  | The time when the condition becomes worse | 5 | 1 | Not provided |
|  | Jaw thrust maneuver | 1 | 0 | Not provided |
|  | Rate of progressing to the critical stage | 1 | 1 | Day 7 |
|  | Time to the critical stage | 1 | 1 | Day 7 |
| Adverse events/effects |  |  |  |  |
|  | Adverse events | 29 | 5 | At baseline, day 3, 6, 10, 14, 28, 90 |

APACHE-II: Acute Physiology and Chronic Health Evaluation; ALT: Alanine aminotransferase; CK: Creatine Kinase; CRP: C-reactive protein; ES: Erythrocyte sedimentation; ECG: electrocardiogram; ICU: Intensive Care Unit; IL: Interleukin; Mb: Myoglobin; NEWS2: National Early Warning Score 2; RNA: ribonucleic acid; SARS-CoV-2: Severe Acute Respiratory Syndrome Coronavirus 2; SOFA: Sequential Organ Failure Assessment.

**Figure 6.**
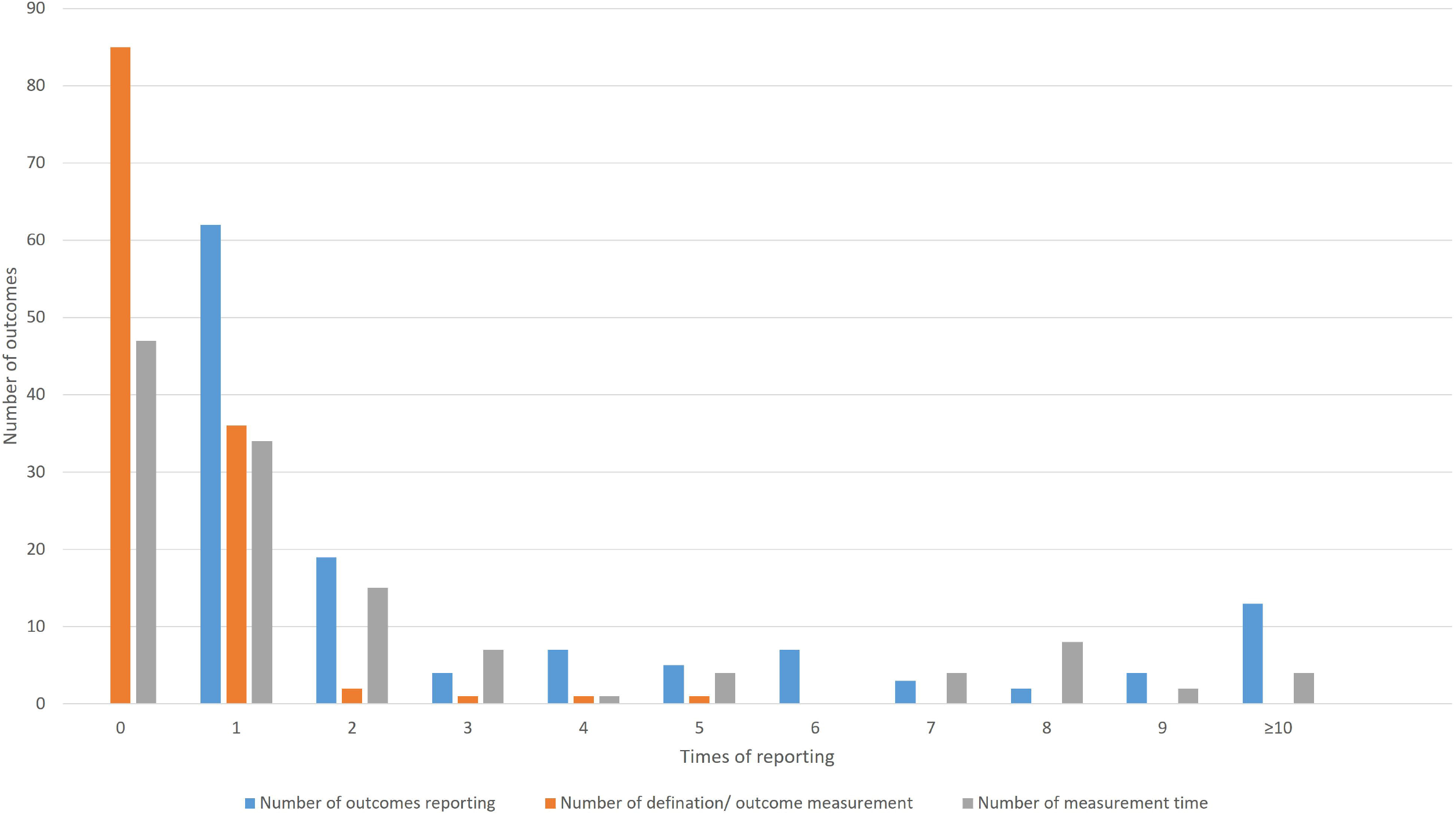
The summary of outcome reporting for protoclas of western medicine clinical trials.

In the 17 outcome domains of protocols of western medicine clinical trials, 5 outcome domains (adverse events/effects, delivery of care, economic, metabolism and nutrition outcomes, mortality/survival) consisted of only one outcome. These outcomes are reported between 1 and 36 times, and the median outcome reporting time is 1. Respiratory, thoracic and mediastinal outcomes included the largest number of outcomes, which includes 31 outcomes; chest imaging is reported more frequently than other outcomes. The number of outcomes in different outcome domains in protocols of western medicine clinical trials is shown in Figure 7.

**Figure 7.**
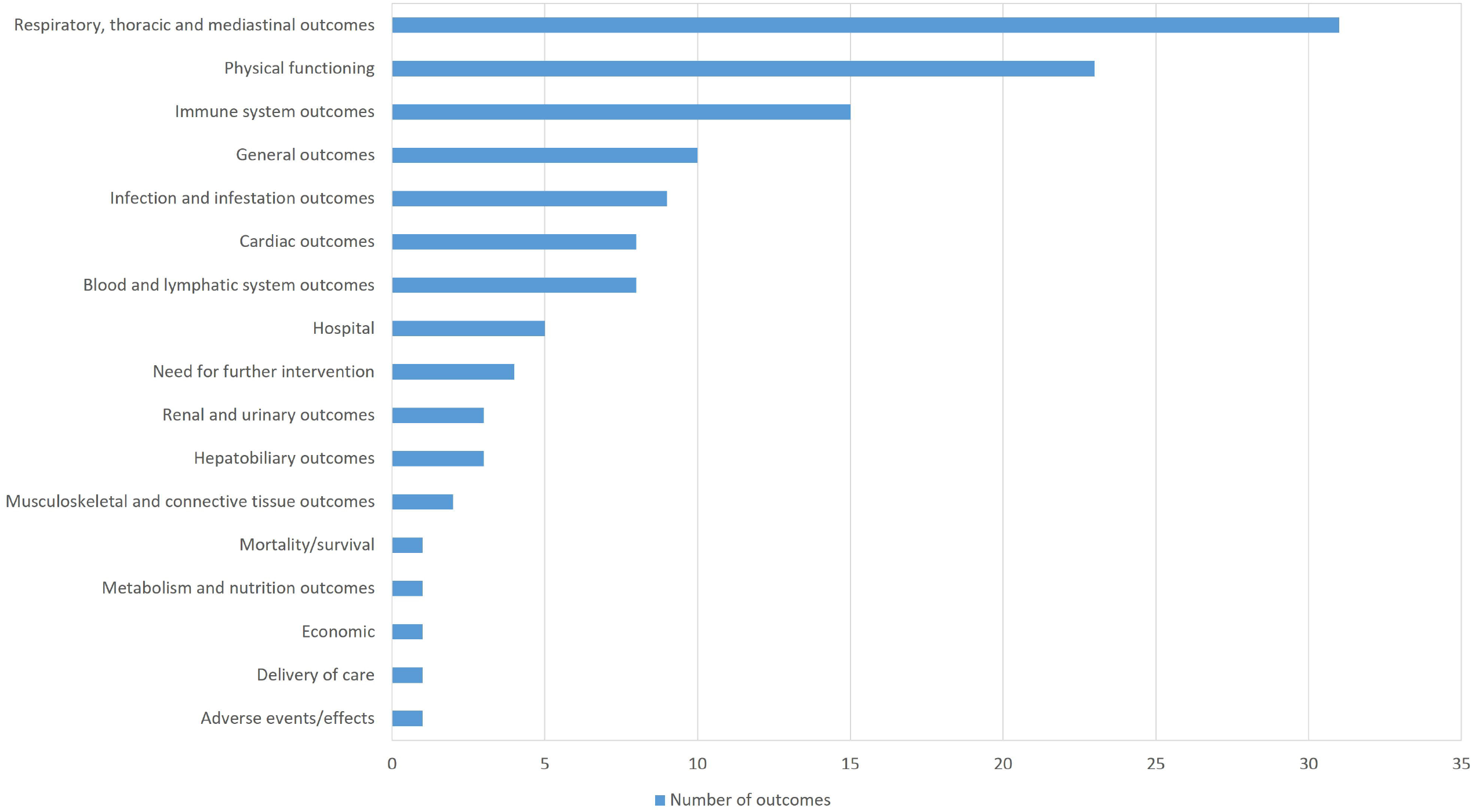
The number of outcomes in different outcome domains in protocols of western medicine clinical trials.

## DISCUSSION

This review is the first to evaluate the outcome reporting of protocols of TCM and western medicine clinical trials for treating COVID-19. The results showed variations in the outcome reporting. For outcome measurement instruments/outcome definitions and outcome measurement time, there is also heterogeneity. However, many primary investigators did not provide outcome measurement instruments/outcome definitions or outcome measurement time. It is difficult to predict results of clinical trials now. But it is obvious that these problems may result in the exclusion of some studies from systematic reviews/meta-analyses due to the heterogeneity of outcomes or outcome measurements. It is a waste.

In this review, we find that there are more than 40 duplicated outcomes between protocols of TCM and western medicine clinical trials. No matter for clinical trials of TCM or western medicine, etiological test, chest imaging, respiratory symptoms, temperature, mortality/survival and adverse events are very important. These outcomes are relevant to the prognosis of disease and safety of therapy.

Because of no specific therapy can be used in the treatment of COVID-19, it is necessary and urgent to conduct clinical trials, no matter what the interventions are. We believe that it is important to develop a COS for clinical trials of TCM and western medicine for treating COVID-19, so that the efficacy of different interventions can be compared and merged in systematic review/meta-analysis.

## Data Availability

The data is from public database and does not include identifiable patient data.

## Contributors

RQ and HS contributed to the study design. XW and MZ conducted searching and extracted data from databases. RQ, CZ, JH, YH, TH contributed to the data analysis. RQ drafted the manuscript. ML, HS, CZ, JC, HS revised the manuscript. All authors read and approved the final manuscript

## Patient consent

Not required.

## Data sharing statement

The data is from public database and does not include identifiable patient data.

